# Making the invisible visible: New perspectives on the intersection of human-environment interactions of clinical teams in intensive care

**DOI:** 10.1101/2021.05.10.21256688

**Authors:** Sheena Visram, Laura Potts, Neil J Sebire, Yvonne Rogers, Emma Broughton, Linda Chigaru, Pratheeban Nambyiah

**Affiliations:** Department of Computer Science/ UCL Interaction Centre, University College London, United Kingdom; Digital Research, Informatics and Virtual Environments (DRIVE) centre, Great Ormond Street Hospital for children, London, United Kingdom; Clinical Simulation Centre, Great Ormond Street Hospital for children, London, United Kingdom

## Abstract

Understanding human behaviour is essential to the adoption practices for new technologies that promote safer care. This requires capturing the detail of clinical workflows to inform the design of new interactions including those with touchless technologies that decipher human-speech, gesture and motion and allow for interactions that are free of contact. Many environments in hospitals are sub-optimally designed, with a poor layout of work surfaces, cumbersome equipment that requires space and effort to manoeuvre, designs that require healthcare staff to reach awkwardly and medical devices that require extensive touch. This suggests there is a need to better understand how they can be designed. Here, we employ a new approach by installing a single 360° camera into a clinical environment to analyse touch patterns and human-environment interactions across a clinical team to recommend design considerations for new technologies with potential to reduce avoidable touch.

## 1 Introduction

There is increasing interest regarding the actions and gestures that healthcare workers use to interact with technology, and the cognitive processes that underpin these interactions. Research to date has predominantly focused on gesture-based control of image manipulation within medical scans,^1^ particularly in operating theatre environments where the demands of sterility restrict the ability of practitioners to use touch-based interfaces.^2,3^ However, many other healthcare environments require numerous human/technology interactions including the Neonatal Intensive Care Unit (NICU).

The complex needs of NICU require simultaneous use of multiple tools and technologies.^4,5^ This need has driven development of miniaturised, wearable and intelligent medical devices ^6,7^ The proliferation of medical devices, monitoring systems, and information technologies in modern healthcare has enabled improved data collection but is associated with reduced standardisation compared to other safety-critical industries^8^ and therefore increased potential for technology-based error. ^9,10^ Human factors and ergonomics (HFE) principles, which arose from post-WWII endeavours to improve the safety of military and transport systems ^11^are important in understanding the relationship between humans and complex technologies and may contribute to healthcare^12^.

The SEIPS (Systems Engineering Initiative for Patient Safety) ^13,14^ approach is one example of a model to support human factors analysis of healthcare systems. A key characteristic of this model involves the description of the work system and its elements (Figure 1). These elements exist within a specific external environment, which may include cultural, regulatory and legal frameworks. Care processes arise from the interaction of these elements, and lead to outcomes for both patients and the organisation^15^. This has led to the design of neonatal incubators, that considers the importance of having clinician control of lighting, sound, access to the infant, alarms and space for the family to bond with the infant ^16^.

**Figure 1.**
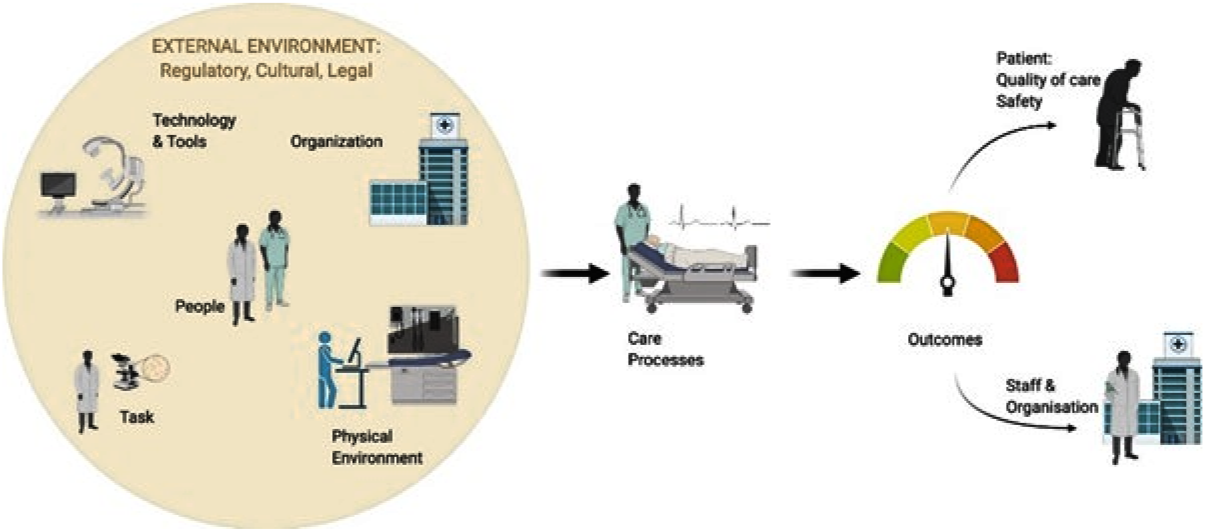
SEIPs system, elements comprise people involved; the technology and tools utilised; the organization; the tasks required of the workers; and the physical environment that these are carried out in. (adapted from Carayon 2014)

Clinical simulation, described as “a technique to replace or amplify real experiences with guided experiences that evoke or replicate substantial aspects of the real world in a fully interactive manner” ^17^ offers an ideal approach to interrogate SEIPS characteristics within common acute scenarios in paediatric healthcare in a safe and controlled setting.

We use a simulated clinical scenario in a safety critical Paediatric healthcare system to develop a coding system for human-environment interactions. The focus of the study was to observe without intervention, the interactions of a clinical team in a NICU environment, using a 360° camera. By taking a behavioural approach to understand touch sequence patterns, behaviours and interactions across a clinical team, the objective was to provide new insights that enhance team performance and thereafter inform touchless technology appropriation and adoption.

## 2 Methodology

A simulated scenario was formulated focused on management of a deteriorating preterm infant; a frequently occurring neonatal emergency. It involves a high number of human-technology interactions common to many intensive care environments resulting in high yield of information for analysis and relevance for a wide range of clinical teams. The simulation session was facilitated by trained simulation faculty according to local guidance. The focus of this exercise was to examine process and interactions rather than deliver training. Participants were therefore briefed about the clinical scenario, the clinical events that would occur and received clear objectives about what clinical tasks needed to be completed. Recruitment was voluntary and participants provided informed consent to participate and be filmed. This study was approved for ethics by University College London under REC ID: 18579.001

A high-fidelity clinical environment was created using a preterm neonatal manikin, simulated monitoring and real medical equipment and devices. The scenario was conducted in real time and was filmed with a small 360° camera mounted on the wall, at a height of 2 metres, fronting on to the cot space. This positioning was selected to give maximum view of the room (Figure 2). There are several 360° cameras available on the consumer market, predominantly developed for extreme sports. The technical specifications of the selected 360° camera were 360° horizontal plane, 240° vertical plane, 4K, 16-megapixel camera and dual microphones. This hardware was evaluated as comparable to its market competitor and was selected due to its 8-element fisheye lens, super wide-angle field of view, lightweight and wipeable lens. Simulation faculty used smartphone controller functions to start, stop and remotely view feed from the camera during filming.

**Figure 2.**
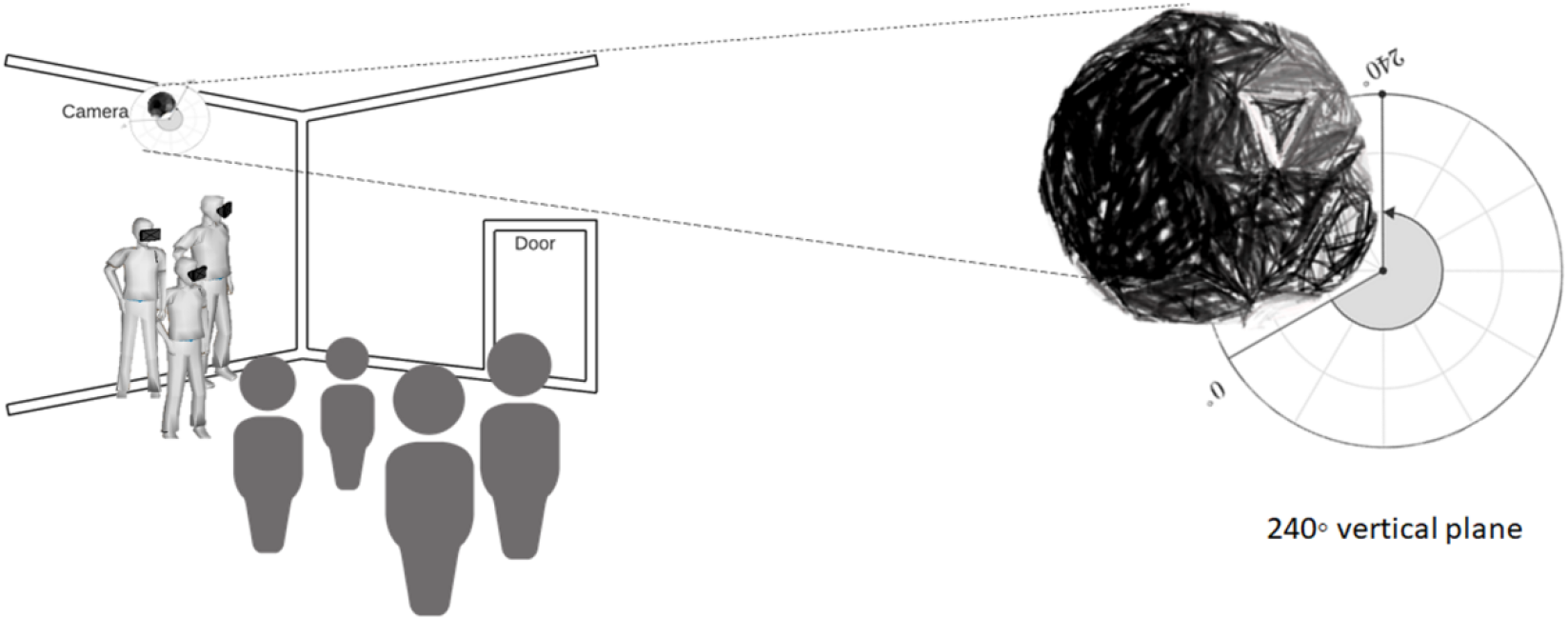
Aerial placement of 360° camera in NICU. This position emulates the aspect of a researcher conducting field observations, with potential analytical gains due to the increased field of view in a single frame from an aerial position to view interactions which might otherwise be obstructed or occluded. This shows how a field researcher would view the clinical workflows wearing a VR headset.

All the clinical objectives of the scenario were met by the team during the scenario. Following the filmed scenario participants engaged in a confidential debriefing discussion, facilitated by simulation faculty and providing opportunities for reflection and discussion and identification of learning points.

Interaction analysis^18^ of the 360° video was completed on a desktop in a 360° viewer, viewing on a VR headset and the audio of the clinical simulation and verbal exchanges were transcribed and matched up to the corresponding tasks observed in the video. Analysis of touch frequency by single touch, sequential touch incidents and patterns of touch across individual team members was conducted by a single analyst by manual review. An episode was defined as a single touch and “a sequence” was identified as a string of two consecutive episodes. Episodes and touch sequence pairs were recounted to check for consistency of the data analysis. Together, these findings were then used to develop a classifier of tasks where behavioural characteristics including avoidable episodes of touch are highlighted to infer the optimal environmental conditions for recognition-based touchless technologies in NICU. These findings were represented as data visualisations to facilitate hypothesis generation on avoidable touch and shared with the participants to develop themes.

## 3 Results

A clinical team who typically work together on a neonatal intensive care unit (three nurses and one doctor who were all female) took part in a clinical simulation lasting 16 minutes and 41 seconds, during which 437 episodes of touch were identified. The simulated scenario identified raised multiple communication, task-based, and environmental considerations.

A surprisingly large number of sequences of touch were identified (n = 68) which make that on average 3-4 a minute. Touch patterns were observed across 17 surface areas and equipment; the incubator (119, 27.2%) and drugs trolley (116, 26.5%) received the highest frequency of touch. A heat map illustrating aggregated episodes of touch by the clinical team during neonatal clinical simulation is available as a supplement. This visually depicts the incubator as a high touch surface area, with a number of touches to the hatches to enter the incubator, the infant as part of assessments and monitoring devices inside the incubator. Of interest, team members touched different surface areas and equipment depending on their roles and responsibilities. Tasks were split across two focal areas; the incubator where Nurse 1 and the Doctor conducted physical examinations and diagnostic tests and the drug trolley, where Nurse 2 and Nurse 3 worked in tandem to prepare medications. Of the 119 touches to the closed incubator, Nurse 1 made 66 (55.5%) of these touches, and of the 116 touches to the drug trolley, Nurse 2 made 77 (66.4%) of these touches. For Nurse 1 this involved tasks that include checking air entry, and vital signs, and also hand offs to receive medications and ancillaries. In a role based at the drug trolley but with frequent interruptions, tasks completed by Nurse 3 involved moving to different parts of the room to source equipment, request for rapid blood tests, cross-check medications being drawn up, set up infusion pumps and an occasion of joining Nurse 1 at the closed incubator to assist in assessing the deterioration.

Touch sequence pairs are displayed using Sankey charting, as a clear graphical visualisation for weighted adjacency data that shows directional ties (Figure 3, Figure 4). By mapping behaviours, to tasks with consideration to touch patterns, we differentiate potentially avoidable touch as: (i) Repeated touch (ii) Obstructions to touch (iii) Unplanned touch (iv) Touch that cause delays. A further category for (v) unavoidable touch is included. This is because it is important to note that episodes of potential avoidable touch occur in amongst several unavoidable touches, which are defined as those that were necessary to perform the sequence of procedures. These observations are used to inform opportunities for touchless technologies (Table 2).

**Figure 3.**
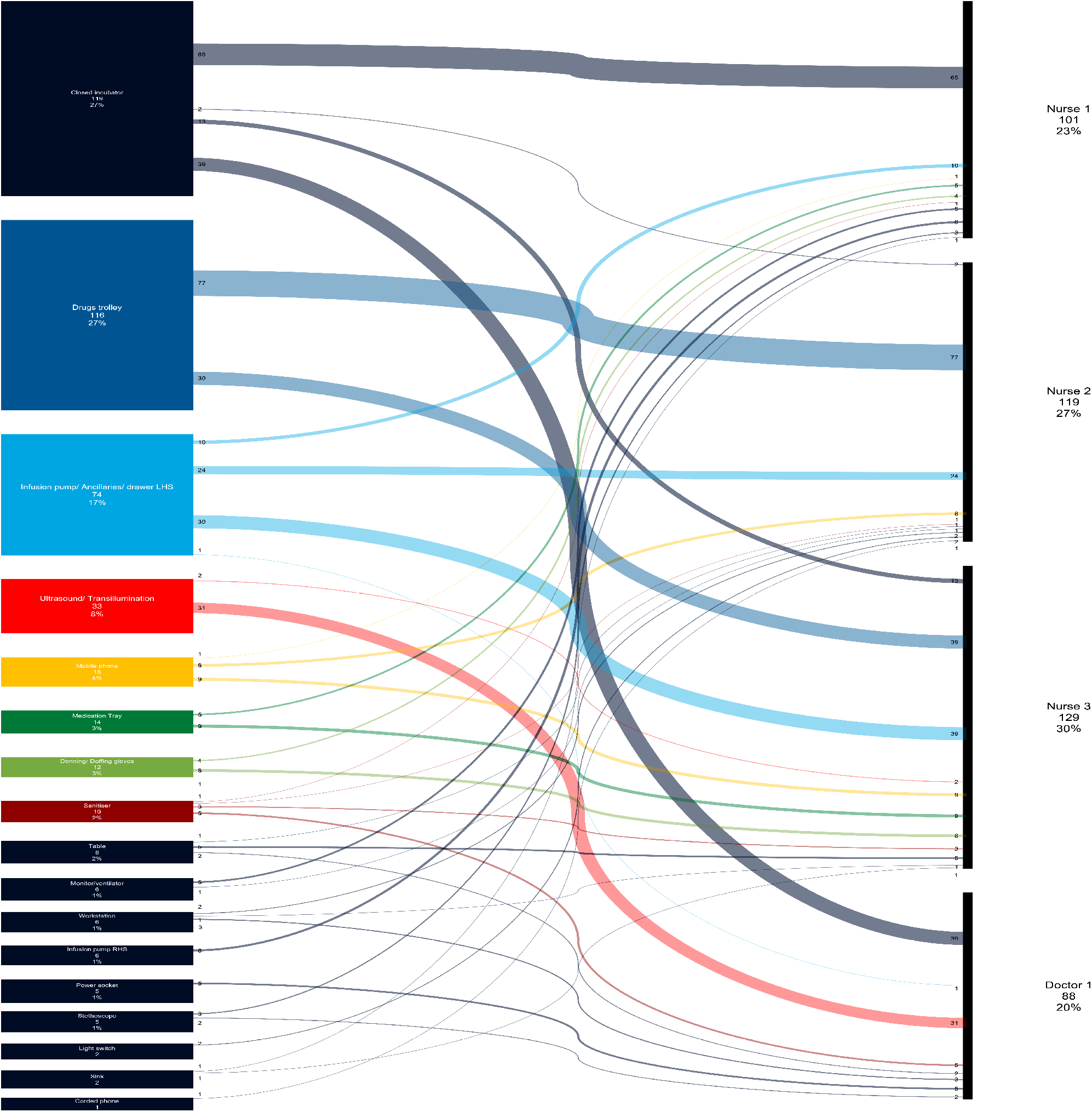
Aggregated touch by equipment and team member accounting for 437 episodes of touch

**Figure 4.**
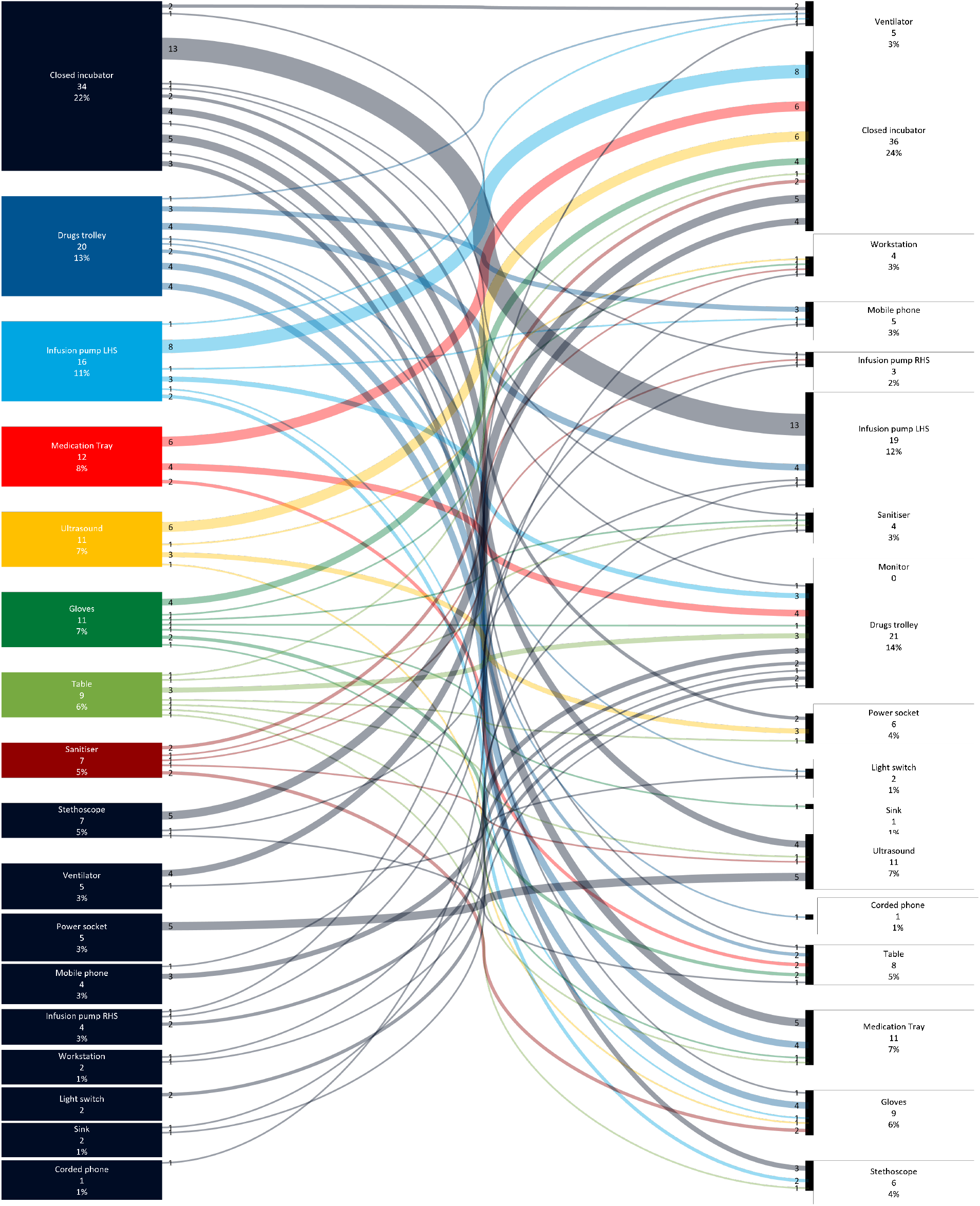
Sub sequences of touch (directed and weighted) in a Sankey chart. The closed incubator, drugs trolley and left-hand sided infusion pump are the top three sources of touch, respectively. The left-hand panel represents the source of a touch sequence pair and the right-hand panel represent the destination of a touch sequence pair.

**Table 1.**
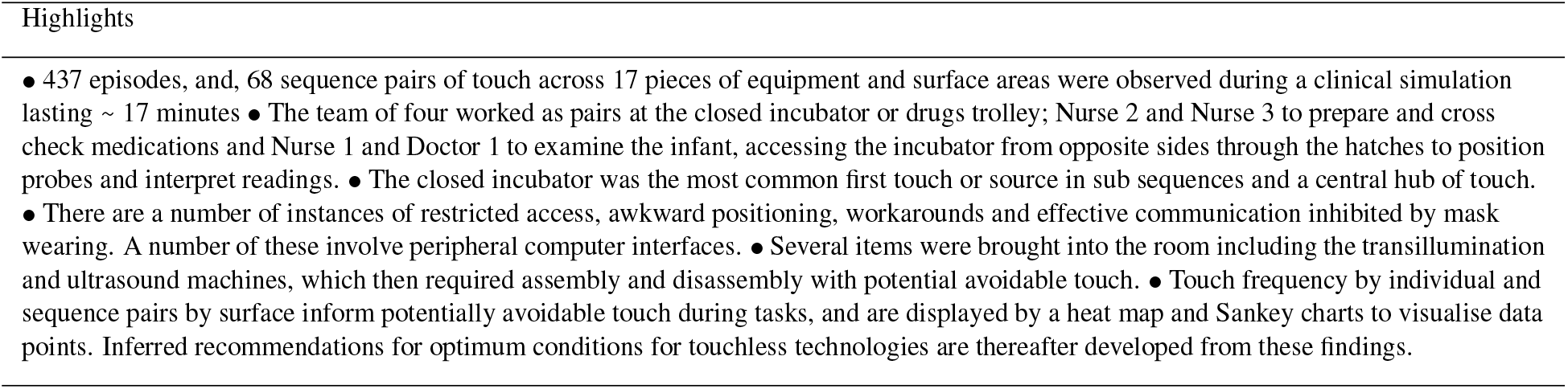
Summary of findings related to touch patterns in NICU

| Highlights |
| --- |
| • 437 episodes, and, 68 sequence pairs of touch across 17 pieces of equipment and surface areas were observed during a clinical simulation lasting ~ 17 minutes<br>• The team of four worked as pairs at the closed incubator or drugs trolley; Nurse 2 and Nurse 3 to prepare and cross check medications and Nurse 1 and Doctor 1 to examine the infant, accessing the incubator from opposite sides through the hatches to position probes and interpret readings.<br>• The closed incubator was the most common first touch or source in sub sequences and a central hub of touch.<br>• There are a number of instances of restricted access, awkward positioning, workarounds and effective communication inhibited by mask wearing. A number of these involve peripheral computer interfaces.<br>• Several items were brought into the room including the transillumination and ultrasound machines, which then required assembly and disassembly with potential avoidable touch.<br>• Touch frequency by individual and sequence pairs by surface inform potentially avoidable touch during tasks, and are displayed by a heat map and Sankey charts to visualise data points. Inferred recommendations for optimum conditions for touchless technologies are thereafter developed from these findings. |

**Table 2.** Mapping avoidable touch to considerations for touchless technologies *EHR refers to Electronic Health Record

| Behavioural mapping to identify potential avoidable touch |  |  |  |  |  | Touchless technology inferences |
| --- | --- | --- | --- | --- | --- | --- |
| Task and team | Repeat touch | Obstruction causing touch | Touch that causes delays | Unplanned touch | Unavoidable touch aspects |  |
| **Checking vital signs**<br>Nurse 1 Nurse 3 | Reaching into incubator.<br>Use of a stethoscope, probe.<br>Multiple touches on wiring. | The incubator door and small hatches for access. Noted constraints of access in and limited space. | Sensors detached inside the incubator that impede assessment. | Repositioned sensors in the incubator | Manual examination of the infant requiring direct touch to complement numerical interpretation of indicators of wellness. | Wireless sensors in the incubator.<br>Monitors activated by voice to alert to results of importance. |
| **Checking air entry**<br>Nurse 1, Nurse 3 Doctor | Suctioning tube and ancillaries passed between team members. | The incubator door and small hatches for access. Noted constraints of access in and limited space. | Hand offs between team to pass, unpack and set up ancillaries. | Stethoscope secured at the edge of the incubator, then placed on the table and moved around the room, needing retrieval. | Manual examination of the infant requiring direct touch to complement numerical interpretation of indicators of wellness. | Wireless smart stethoscope located within the incubator. |
| **Setting up infusion pump**<br>Nurse 3, Nurse 2 | Multiple touches to set rate and volume of pump. | Placement of infusion pump by side of incubator resulting in pinch point to access. | Multiple alarms which delay infusion. | Multiple alarms which interrupt other tasks. | The design of the infusion pump does not allow for touch free interactions or interoperability with EHR | EHR interoperable infusion pumps, to set rates, monitor volumes and record prescription times and dates. |
| **Drawing up medication**<br>Nurse 2 Nurse 1 | Multiple touches on drug trolley, unmarked drawers inside and outside room. Multiple touch to mobile phones for calculations. | Draws are closed, unmarked and opaque and do not allow overhead view of medications, syringes and ancillaries before opening. | Two people required to cross check and administer medications. | Multiple draws of ancillaries located outside room. | Medications needed to be drawn up into syringes manually and cross-checking medications is a safety step. | A table-top or wall mounted screen for access to inventory and to replace mobile phone use. |

Table 2 continued
|  |  |  |  |  |  |  |
| --- | --- | --- | --- | --- | --- | --- |
| **Examine** Doc-<br>tor 1, Nurse 2 | Repeated touch between ultrasound/transillumination and incubator. | The incubator door and small hatches for access. Noted constraints of access in and limited space. | Limited in room equipment for investigation. Numerous exits from the room to retrieve equipment. | Assembly of new equipment, unravelling cables, and accessing power points. Manual switch to adjust lighting. | Direct touch is required to examine and orientate probes over infant as a safety step. | Wireless ultrasound probes. Perspex over incubator to display vital signs or project interactive information in real time including ultrasound or x-rays. |
| **Entry into medical records** Doc-<br>tor 1 | Workstation with desktop, keyboard, and mouse. Mouse used to navigate EHR*, multiple clicks for navigation. | The workstation is positioned with the screen obstructing the view of the incubator. | Stretching to see monitor at other side of room to be able to enter data, confirmation of values from other team members. | Some equipment (i.e ultrasound) utilise new monitors which add new temporary focal and touch points. | The workstation interface and EHR* is not interoperable with data collected by devices in the incubator. | Pre-programmed smart phrases and gestures. Interoperability of the EHR with IoT devices in the room as part of a local digital health ecosystem |

**Table 3.** SLOAN framework: Optimal conditions for touchless technologies by space, lighting, obstructions, and ambient noise in NICU created from observations made during the interaction analysis.

| **Space** | **Lighting** | **Obstruction** | **Ambient Noise** |
| --- | --- | --- | --- |
| **Protected space:** Any new technology cannot interrupt route of flow of ‘walk around space’ for incubator, drugs trolley as prime focal points or hygiene-promoting facilities like the sink or sanitiser<br>**Integration:** Any new technology must be integrated with the most relevant equipment (like the NICU bed space, EHR, PACS**) and not require a process of retrieval, assembly, installation, or personalisation.<br>**Placement:** Any new technology must not require a table or secondary surface for placement and set up. The surface of the new technology must be easily cleanable. | **Light adjustment:** Lighting adjustments for the needs of the new technology should be positioned within reach without interruption from other tasks.<br>**Focused lighting:** * Multi – level lighting for illumination for a new technology should be directed away from the incubator<br>**Low level lighting** New technologies that are recognised in low levels lighting would be optimal | **Back-to-back Communications:** Any new technology should not proliferate obstructions to effective communication such as back-to-back verbal exchanges amongst the team<br>**Distraction:** Any new technology must not obstruct or distract attention from the clinical workflow and prime focal points of care but rather be glanceable and easily interpreted<br>**Access:** Any new technology must overcome constraints and obstruction of access into and around the incubator space. | **Pitch:** * Any new technology must not introduce noise beyond recommended levels or introduce new systems that alarm<br>**Personal protective equipment:** Any new technology must be compatible with wearing personal protective equipment like masks, gloves and gowns. Recognition based software should be sensitive to the constraints posed by this.<br>**Cross talking:** Any new technology must be able to differentiate commands from team members without complicating the flow of verbal exchange |

\*Derived from White <sup>19,20</sup>
\*\*PACS refers to Picture archiving and communications system

A series of conditions are then hypothesised to facilitate the successful integration of a given recognition based touchless technology or sensor in NICU. Inferences for optimum conditions for touchless technologies are based upon observed characteristics of the physical environment and sensory modalities of light and sound.

When combined with findings from the interaction analysis, these are collated as design recommendations and a 12-point SLOAN framework. This is intended as a quick reference tool for use by human computer interaction (HCI) teams to drive cooperative design thinking with clinical teams, and, for developers creating touchless technologies.

## 4 Discussion

This study of a simulated acute clinical scenario in NICU showed many sequences of touches from person to equipment to technology that potentially could be reduced, and workflows that could be improved by configuring the space differently. The analysis identified touch frequency and patterns for analysing human-environment interactions to appropriate technologies that have the potential to reduce touch.

The seminal US Institute of Medicine report To Err is Human was a tipping point for patient safety activity^21^ and cognitive psychology and human factors engineering methods were foregrounded in the discussion of patient safety. In the decades since, healthcare has become more advanced but also more fragmented and decentralised. Attempts at improving safety often focus on behavioural change of individual healthcare workers, either because these are perceived to be the most important factors, or because they are perceived as quick wins. However, it has been recognised that attempts at behaviour-centric change often fail without concomitant system change^11^. The SEIPS framework describes key characteristics of a model which seeks to address such systems factors. The current findings demonstrate that touch is a fundamental part of healthcare activity, forming the basis of the relationship between healthcare workers, their tasks, tools, and the physical environment they work in. Intensive care environments are complex and involve a high incidence of human-technology interactions.

This is the first observational study to our knowledge that has focused on the translation of hand touch patterns across a clinical team during clinical interactions to identify potentially avoidable touch with the intention of inferring opportunities for touchless technology implementation. Reducing avoidable touch could contribute to reducing the number of high touch surfaces including peripheral computer interfaces, that may improve patient safety by reducing the hospital acquired infections associated to this ^12,22,23^. There are additional efficiency and protective gains for healthcare workers who would be required to don and doff PPE less frequently^24^.

New technologies that might be considered include wireless sensors in the incubator, the use of voice and hand gesture recognition technologies to interact with peripheral computer interfaces and use of the Perspex screen over the incubator to display real time status updates about the infant. Each of these also have the potential to reduce episodes of touch within the NICU environment. Importantly, interoperable exchanges of health information are considered with the potential to reduce duplication and make available the information collected by a new touchless technology as part of the EHR.

This study of a simulated acute clinical scenario in NICU identified touch frequency and patterns for analysing human-environment interactions to appropriate technologies that have the potential to reduce touch. Surfaces are touched by different team members, working as pairs and as part of tasks that make up clinical workflows. Of the tasks completed during workflows, four types of potential avoidable touch were analysed juxtaposed with unavoidable touches for care provision. This novel perspective on human-environment interactions highlights that the focus of identifying avoidable touch and the role of touchless technology solutions must be considered in the context of characteristics of team behaviours and environmental interactions that make up clinical workflows and not simply touch patterns. NICU is complicated by frequent flows of decisions and that to be useful in NICU, new technologies not only need to match the reliability of traditional methods, but also should be able to add valuable information to the clinical team as part of a clinical workflow and hence form an integrated solution^25^. Although outside the immediate scope of this analysis, this study also proposes design recommendations for the effective use of space, suitable lighting, minimising obstructions and limiting ambient noise to improve integration of new interactions with touchless technologies with the goal of improving safety, efficiency and wellbeing.

This preliminary study observes a high-fidelity simulation of the heterogenous, real time environment of NICU conducted during the COVID-19 pandemic. Even outside of pandemic restrictions, there are few qualitative studies of design considerations in NICU. This is because these studies typically require extensive on-site observations and interviews^12,26^. By contrast, this study was interested in less invasive and remote methods of evaluating interaction design with computer interfaces in hard to reach, safety critical environments. To collect data, a 360 camera was used that enabled clinical workflows to be observed through recreating the simulation in a real-life virtual environment. In particular, the use of the 360° camera allowed an enriched, immersive and empathetic researcher experience when reviewing a predetermined timeline. It provided a vantage point to view a wider aspect and angle of the room, view simultaneous tasks in different areas that would otherwise be missed and to do so without interruption to the task at hand. This analysis offers new insight into the workings of a clinical team, in an environment with robust and evolving design guidelines by using human-centred design strategies to highlight potential targets for new touchless technologies^27,28^. Similarly, there are several hard to access or yet to exist places that 360 films of live spaces or design concepts can be used to evaluate. By using a new approach, both of an unobtrusive 360° camera, and touch pattern analysis this could act as a paradigm for multiple novel future evaluations of environments in healthcare in a safe and remote way.

## Data Availability

Supporting information and supporting data are available from the authors upon request.

## 5 Authors’ contributions

SV, PN conceived the project. LP, EB, LC were involved with the simulation design. LP led the simulation session and filming. SV provided the camera equipment, conducted the analysis and reported on findings creating Figures 2-4 and Tables 1-3. NJS, YR provided supervisory oversight of the project and revisions on early drafts of the manuscript. All authors read and approved the final manuscript.

